# Seasonal and regional heterogeneities in antibiotic resistance (AbR) across meatpacking plants in the USA

**DOI:** 10.64898/2026.01.12.26343976

**Authors:** K. W. Okamoto, R. G. Wallace, A. Liebman, H. G. Peterson, C. Freshour, A. R. de Campos Silva

**Affiliations:** Department of Biology, University of St. Thomas, St. Paul MN 55105 USA; Agroecology and Rural Economics Research Corps, St. Paul MN 55105 USA; Department of Geography, Rutgers University, Piscataway, NJ 08854 USA; Regions Refocus, New York, New York 10001 USA; Department of Veterinary Clinical Sciences, College of Veterinary Medicine, University of Minnesota, St Paul MN 55108 USA; Department of Geosciences, Georgia State University, Atlanta, GA 30303 USA; Instituto de Políticas Públicas e Relações Internacionais, Universidad Estatal Paulista, São Paulo, SP CEP 01001-900 Brazil

## Abstract

A foundational question in public health is why certain infectious diseases emerge in some locales at certain moments, but not others. Here we leverage a multiyear dataset on nearly all meatpacking plants in the USA handling chicken, cattle, swine, and turkey products as a case study to present a novel approach to elucidating the spatiotemporal dynamics of a major human disease agent: antibiotic resistant (AbR) bacteria. In particular, we apply geographical and temporal neural-network weighted regression, a novel spatiotemporal analysis based on deep-learning, to characterize how infection risks at discrete facilities shift over space and time within the United States. Bracketing the dataset from 2021-2024 by seasons and processing plants across the US, our analyses find seasonal and regional differences in AbR bacteria detection by plant size and commodity type. These factors differentially affected when and where AbR bacteria were found — from distinct outbreaks specific in time and place to broader and persistent trends. While highlighting environmental and occupational safety hazards posed by AbR bacteria, our analyses have the potential to help local communities within plant environs and beyond to operationalize interventions into epidemiological risk profiles posed by dynamic commodity production regimes across spatial scales.

## Introduction

It is hard to overstate the centrality of spatiotemporal heterogeneity to public health (Elliott & Wartenberg, 2004; Thomas et al., 2020). From proximity to carcinogens such as airborne particulate matters (Hamra et al., 2014; Wang et al., 2025) and industrial toxins to fire hazards (Wallace & Wallace, 2011), disparities in access to resources such as clean drinking water (Rosinger & Young, 2020), family-practice clinics (Naylor et al., 2019), well-designed sidewalks (McCormack & Shiell, 2011; Omura et al., 2020), and climatic clines in pathogen and vector population densities (Pecl et al., 2017), almost all health exposure risks exhibit distinct geospatial signatures over time.

Antibiotic resistant (AbR) bacterial infections are no exception (Baym et al., 2016). Antibiotics work by either killing or preventing the reproduction of bacteria cells (Kohanski et al., 2010). Yet bacteria populations frequently evolve to mitigate such adverse fitness effects, resulting in compounds that previously inhibited bacterial population growth no longer able to do so effectively (Davies & Davies, 2010; Blair et al., 2015; Munita & Arias, 2016). The subsequent spread of mutations or novel genes facilitating AbR within and across bacteria populations is an inherently spatiotemporal phenomenon (Baym et al., 2016; Okamoto et al., 2018).

The public health consequences of antibiotic resistance (AbR) have received considerable attention (reviewed in Davies & Davies, 2010; Laxminarayan et al., 2013; Holmes et al., 2016; Elshobary et al. 2025) and for good reason. Some estimates attribute over 1.27 million annual fatalities worldwide to drug-resistant bacterial infections and the total global disease burden of AbR has been estimated at ∼47.9 million disability-adjusted life years (DALYs) (Mohsen Naghavi, 2022; Kim et al., 2023; World Health Organization, 2023). As AbR spreads across bacterial species and populations, these numbers are projected to rise considerably in the coming decades (Tiseo et al., 2020; GBD 2021; Antimicrobial Resistance Collaborators, 2024). Nevertheless, there is surprisingly little consensus on the proximate processes driving the origin and early dissemination of widespread AbR. Among many hypotheses, intensive antibiotic use in agriculture has earned much attention (Udikovic-Kolic et al., 2014; Van Boeckel et al., 2015; McKenna, 2017; Irfan et al., 2022; Mulchandani et al., 2023). The rate at which AbR evolves in bacterial populations depends in part on the level of antibiotics to which a bacterial population is exposed (Drlica & Zhao, 2007; Gullberg et al., 2011; Andersson & Hughes, 2012; Blake et al. 2025). Indeed, according to the FDA Center for Veterinary Medicine, about two-thirds of the medically important antimicrobial drugs sold in the United States are intended for use in food-producing animals (FDA CVM, 2024a,b). Agriculture’s relatively heavy reliance on antibiotics has led some investigators to propose that food production plays a notable role in selecting for AbR bacteria of clinical concern (Landers et al., 2012; Irfan et al., 2022; Acosta et al., 2025).

Whilst strong selection pressure for AbR may exist at the farm or ranch level, how AbR mutations that occur in such settings might subsequently disseminate to eventually present in human patients in the clinic remains poorly understood (Verraes et al., 2013; Founou et al., 2016; FDA CVM, 2024a). Of particular relevance is the fate of AbR mutations at highly interlinked supply chain junctures (Xu et al., 2025). Because they bridge sites potentially selecting for AbR emergence in the first place to sites where human AbR exposure risks occur, characterizing spatiotemporal patterns of AbR at intermediary steps of the supply chain can provide unique perspectives on how selection for, and infection by, AbR bacteria ultimately connect (Innes et al., 2021, 2023; FDA CVM, 2024a). As major intermediaries between a range of specialized animal husbandry operations and regional, end consumer-facing retailers handling a variety of commodities, meatpacking plants play an integral role in both facilitating, but also meaningfully mitigating, the spread of AbR that may emerge and evolve to selection pressures at the farm or ranch level (Savin et al., 2021; Hamilton et al., 2024; Yang et al., 2024). In addition to providing crucial insight into how AbR bacteria spread from putative points of selection to infection, characterizing how the distribution of AbR at such facilities changes over time across large spatial scales can provide unique insights into the changing biogeography of AbR exposure risks (Visvalingam et al., 2016; Cobo-Díaz et al., 2021; Innes et al., 2023; Pham et al., 2025).

To this end, we propose a strategy to assess the spatiotemporal distribution of AbR in select supply-chain nodes across several seasons on a continental scale. We illustrate our approach integrating data from a publicly available database from the United States Department of Agriculture (USDA). Our strategy employs a variation on a recently proposed geostatistical method based on deep learning — namely, geographically- and temporally-weighted neural network regression (GTWNNR) (Wu et al., 2021; Yin et al., 2024) — to analyze how the effects of facility scale and commodity type on AbR detection potentially vary over time and space. This environmentally heterogenous dataset enables investigating how the secular combination of regional and seasonal drivers can interact with supply-chain specific variables in affecting AbR detection levels. The proposed framework enables researchers to begin to assess when, where, and how differences between meatpacking plants may matter with respect to AbR exposure risks. More generally, our approach enables epidemiologists to begin to interrogate how health risks are embedded within extant supply-chain processes can cascade over space and time to impact localized health risks (Clark et al., 2025).

## Materials and Methods

### Data

We use as the basis of our analysis datasets from the USDA’s Food Safety Inspection Service (FSIS). For several reasons, but at least in part out of recognition of their central role in linking potential farm-level AbR selection pressures to exposure risks to the broader population, the FSIS routinely monitors meatpacking plant operations throughout the United States, including its territories and colonies, for the presence of AbR bacteria (CDC, 2024; FDA CVM, 2025; USDA FSIS, 2025b).Sampling occurs at regular intervals at the facility level, with samples tested for the presence of contamination by a major bacteria pathogen (generic *Escherichia coli*, *E*. *coli* O157:H7, Shiga toxin-producing *E*. *coli*, *Salmonella* spp., *Campylobacter* spp., *Listeria* spp. and *Enterococcus* spp.; USDA FSIS, 2025c; FDA CVM, 2025). As of 2025, contaminated samples are subsequently evaluated via genetic sequencing and laboratory cultures for antibiotic resistance (for the full list of antibiotics screened, see USDA FSIS, 2025a).

Further details on facilities are conveyed in the dataset. For instance, for each facility its production scale, the date of sample collection, the commodity handled, geographical location, among other details, are demarcated. The data available on the FSIS website (USDA FSIS, 2025c) cover select major livestock species in the US we used for our analyses: cattle, swine, chicken, and turkey.

Although the earliest records in the datasets date back over a decade, we focus our analysis on the three-year period between Sept. 1 2021 and Sept. 1 2024 for several reasons. First, not all species are present in the data since 2014, when the earliest data are available, making comparisons across commodities challenging. Second, since 2013 the USDA has called for phasing out the use of antibiotics for non-therapeutic purposes in livestock production (FDA CVM, 2013, 2019). Thus, early records may reflect far from uniform patterns of antibiotic use at the farm level that would be challenging to interpret with meatpacking plant data alone. By using later data, when presumably more livestock operations have aligned with the USDA’s request to use substantively fewer antibiotics (FDA CVM, 2019), we can potentially reduce the likely confounding effect of dramatically differing farm-level compliance, and hence selection pressures on AbR. Finally, as has become quite well-documented, at the onset of the 2019 SARS-CoV-2 pandemic, the unique, localized operating conditions that persisted at meatpacking plants resulted in substantial disruptions to the supply chains of major meat products (Waltenburg, 2020; Whitehead & Kim, 2022; Luxton & Malin, 2025). While potentially highly insightful, pooling data before and after the early years of the SARS-CoV-2 pandemic risks obscuring broader spatiotemporal trends. For instance, locales varied considerably in their response to the early outbreaks — responses which also varied markedly over time (Davis et al., 2011; Taylor et al., 2020). By focusing our analyses on AbR data since Sept. 2021 — by the time major vaccines against SARS-CoV-2 became widely available in our study regions — we aim to obtain at least somewhat greater clarity on spatiotemporal trends across meatpacking facilities under potentially more comparable conditions over time. Over the time horizon we analyzed, sufficient data were available to conduct our analyses on facilities from the 50 US states, Puerto Rico, and the District of Columbia.

### Analyses

We evaluate how AbR detection across facilities is driven by the interplay of three proposed drivers: regional processes operating at local spatial scales, changes in local AbR prevalence over time, and spatial characteristics of the meatpacking plants themselves. We build on geographical and temporal neural network weighted regression (GTNNWR) as an analytic framework to characterize how these three drivers of AbR differences potentially interact (Fig 1A). The basic idea behind GTNNWR can best be described by beginning with a characterization of classic geographically weighted regression (GWR)( Brunsdon et al., 1996). As in traditional multivariate regression, a response variable *Y* is assumed to depend linearly on potential explanatory variables *X*_1_, *X*_2_, . . . *X_n_* with weights *β_xi_* associated with each explanatory variable *x_i_* (Hastie et al., 2009; James et al., 2021). In GWR, the weights or effect sizes *β_xi_*(·) of explanatory variable *x_i_* are modeled to be specific to each spatial location (typically represented as latitudinal and longitudinal coordinates), and thus the effect of the explanatory variable *x_i_* potentially differs across sites (Brunsdon et al., 1996; Fotheringham et al., 2002). Geographically and temporally weighted regression (GTWR) is a natural extension of GWR where the effects of variable *x_i_* also vary over time. The basic regression formula for GTWR is given by:

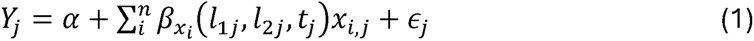

**Figure 1.**
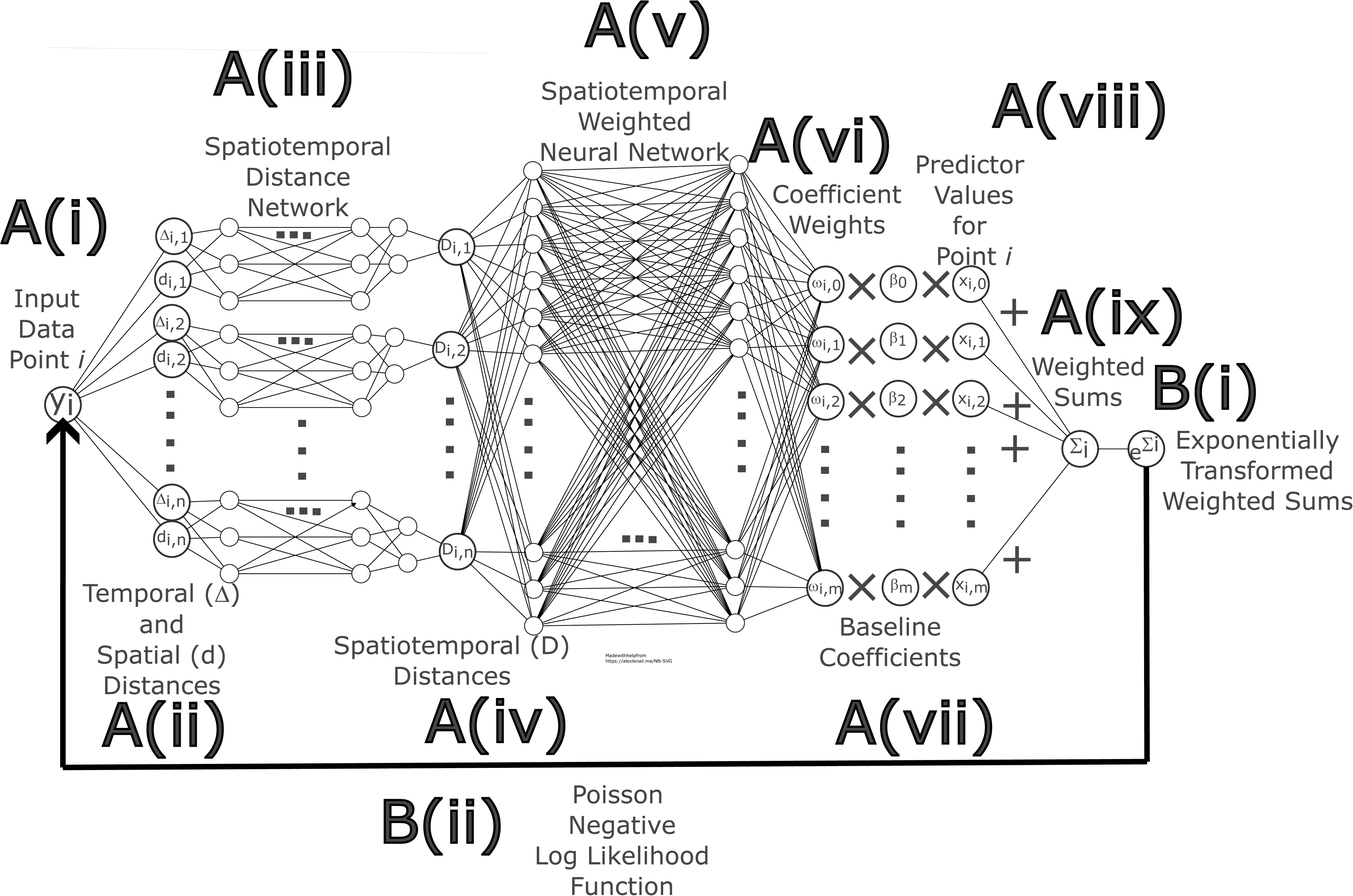
A schematic illustration of (A) the geographic and temporal neural network weighted regression fitting regime (GTNNWR), after Wu et al. 2021, and (B) the modifications we propose to handle count data using a Poisson negative log likelihood (NLL) loss function (a Poisson GTNNWR or pGTNNWR). In its original formulation, a GTNNWR is fitted after partitioning the dataset into training, testing and validation data at random. (A(i)) For each point in the training data, (A(ii)) the temporal (Δ) and spatial (*d*) distances are calculated between that point and all *n* other datapoints. (A(iii)) A spatiotemporal distance neural network is then fitted with the using the temporal and spatial distances and the input layers, and, for each pair of data points, (A(iv)) an output layer consisting of the combined spatiotemporal distances is obtained. (A(v)) A second spatiotemporally weighted neural network is then constructed to predict coefficient weights for each of the *m* predictors (A(vi)), which are then scaled by the baseline coefficients (A(vii)), including the intercept. The baseline coefficients can potentially be estimated, for example, with ordinary least squares or, alternatively, assigned as constants. The regression coefficients are then multiplied by the *m* predictors for each datapoint, and their sum (A(ix)) becomes the provisionally predicted value. In a pGTNNWR, (B(i)) this sum is then transformed using an exponential function, and (B(ii)) the performance evaluated via a Poisson NLL loss function. The regression model is then iteratively fit on resampled training data and adjusting the parameters of the neural networks to minimize the loss function.

where *Y_j_*represents the value of the response variable of the sample *j*, *l*_1*j*_,*l*_2*j*_ represent the spatial position of sample *j* (typically expressed as latitude and longitude), *t_j_* describes the time at which sample *j*’s data were obtained, *x_i,j_* represents the value of the *i^j^* explanatory variable for sample *j*, and *ϵ_j_* describes the random deviation of the *j^th^* record’s response value from its predicted value (Huang et al., 2010).

A key step in fitting a GTWR is specifying the spatiotemporal kernel, which weighs how localized effects decay over space and time for a particular explanatory variable. In essence, GTWNNR differs from classical GTWR by characterizing the spatiotemporal dependency among samples basically using a neural network as a kernel rather than traditional kernel functions such as Gaussian or bisquare representations (e.g., Fotheringham et al., 2017; Fig. 1A). The output layer of the neural network — which accounts for the effects of spatiotemporal distances between samples — is then used to weigh the effect coefficients *·*.. The response variable’s value is then predicted as a linear combination of the weighted effect coefficients and the explanatory variables. The fit is then measured using a loss function, and the parameters of the neural network are then iteratively adjusted until the value of the loss function is minimized. Typically, further iterative refinement occurs on training data, and the GTNNWR’s performance can then be evaluated against validation and testing data.

Here we build on this basic approach to deal with very common types of epidemiological data. Existing implementations of GTNNWR assume the response variable *Y_j_*to take on continuous values. This, in turn, suggests adopting loss functions with attractive mathematical properties to fit the neural network, such as the Mean Squared Error (MSE). However, as with many epidemiological datasets, our response data consist of discrete counts (i.e., the number of AbR bacteria samples detected during inspections). Hence, we take a further step that broadens the application of GTNNWR described above to such data. In particular, prior to evaluating the adequacy of the GTNNWR’s predicted fit on a given iteration, we transform the predicted response variable 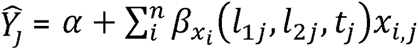 with an exponential function to ensure the fitted values are positive. We then apply a Poisson negative log likelihood (NLL) loss function (Paszke et al., 2019) to characterize the resulting fit (Fig 1B). A Poisson NLL loss function is often used in the neural network literature when the target — here, our response variable, i.e., the number of AbR positive samples — consists of count data (Terven et al., 2025). For convenience, we refer to our approach (Fig. 1B) as Poisson GTNNWR, or pGTNNWR to distinguish it from classic GTNNWR modeling (Fig 1) (Wu et al., 2021; Feng et al., 2021; Hagenauer & Helbich, 2022; Yin et al., 2024).

Our proposed basic strategy for pGTNNWR is in part inspired by Poisson deep neural network (PDNN)— modeling, an approach that has been successfully applied to predict heterogeneities in disease occurrence (Montesinos-López et al., 2020). In particular, Montesinos-Lopez et al. (2021) used PDNN modeling to predict the genetic basis of wheat (*Triticum aestivum* L.) resistance to the fungal pathogen *Fusarium graminearum*. In their study, a PDNN evaluated how single nucleotide polymorphisms (SNPs) interacted with growing conditions to predict individual wheat plant disease severity, measured as counts of infected spikelets on each stalk (Montesinos-Lopez et al., 2021). The predictive abilities of PDNN outperformed several conventionally used inference strategies (including traditional generalized Poisson regression and an array of Bayesian ridge regressions), although classical deep neural network modeling relying on continuous approximations to count data also performed competitively (Montesinos-Lopez et al., 2021). Thus, PDNN models’ relative success in predicting the genetic basis of disease severity suggests the strategy we propose for applying pGTNNWR to count data combining an exponential transformation of the output layer and Poisson NLL function may be a promising approach to elucidating spatiotemporal heterogeneities in the emergence of infectious pathogens such as AbR bacteria.

### Application

To characterize spatiotemporal heterogeneities in AbR, we identified the number of positive AbR bacteria samples for each meatpacking facility in the dataset at a given moment in time. For our analyses, we used meteorological seasons (starting on September 2021 or the beginning of meteorological autumn 2021 and ending in November 2024 or the end of meteorological autumn 2024) as the relevant time unit. This decision on the temporal unit of analysis was based on several considerations. First, and perhaps most notably, much of the US is located between temperate latitudes. Hence, there can be considerable climatic variability over time in growth conditions experienced by bacteria. This temporal variability, particularly with respect to temperature, humidity and rainfall, can be well-characterized by local meteorological seasons (NOAA, 2016). Relatedly, the conditions under which livestock are raised (e.g., Allen Tucker, 1982; Miao et al., 2005; Mendes et al., 2012; Wu et al., 2019; Apalowo et al., 2024; Oke et al., 2024), transported (e.g., Ellis & Ritter, 2006; Watts et al., 2011), and ultimately processed, and thus the potential selective pressures for AbR and its detectability, may also reflect climactic, rather than simply calendar, temporal differences. To be sure, animal production and its potential impacts on AbR can often be less a function of the natural economy of birthing. For instance, apart from key holidays — e.g., turkey for Thanksgiving in the US — the inherent biologies of a laying or birthing season can be circumvented by continuous intensive production under artificial lighting regimes (e.g., Sykes, 1968; Olaf Thieme & Dafydd Pilling, 2008; USDA Animal and Plant Health Inspection Service, 2013; Fuks et al., 2022; Ferreira et al., 2024; Sutherland et al., 2024; Fairchild, 2025). Whether bacteria themselves are similarly detemporized in these systems remains an open question. Despite this potential caveat, in our view, using meteorological seasons allows us to assess how sources of environmental variation potentially operates across different geographic regions to impact AbR prevalence.

A further consideration in our choice of temporal unit is dictated by the nature of the data collection regime. FSIS microbiological surveillance is conducted on structured, recurring schedules in which slaughter establishments are assigned a defined number of sampling tasks per month based on slaughter volume under the NARMS program, with the largest facilities sampled at a frequency of approximately once per week, rather than through continuous daily collection (USDA FSIS, 2021b, 2024). This leads to unbalanced presentation of sites, regions, and predictors in the dataset over too fine a temporal scale (e.g., day or even week). By integrating sample outcomes for a meteorological season, we can aim for greater consistency in the number of times each site appears in the dataset per time unit. Across all seasons, each site appears at least once per season, and the median number of records for each site per season is 2, with a mean of 4.85 and standard deviation of 4.92. To be sure, our approach using meteorological seasons to demarcate subsets of data meant there were a small number of gaps for a few locations in some seasons. Nevertheless, the vast majority of plants were consistently captured at this unit of temporal resolution throughout the time horizon we investigated. Hence, between both the data-specific and biological considerations described above, we believe meteorological season emerges as a potentially informative temporal unit for our analyses.

To further describe AbR incidence in meat processing plants across the US between late 2021 through summer 2024, we use pGTNNWR to characterize how processing plant size and commodity type might affect AbR detection over space and time. The dataset divides the processing plant sizes into three categories: large, small, and very small, and ten commodity types are used to characterize the product each plant handles (Table 1).

**Table 1.** Variables used as predictors for our Poisson Geographical and Temporal Neural Network Weighted Regression (pGTNNWR), and the number *N* of facilities fulfilling those criteria in the dataset. Note that for commodities sampled, a given processing plant can potentially handle multiple products. We add that for the product “Animal - young chicken carcasses”, another commodity present in the dataset, the number of records was too small (*N*=3) to enable reliably including it in the analyses.

| Variable | Values | Products, if applicable | $N$ |
| --- | --- | --- | --- |
| Processing Plant Size | Large |  | 276 |
|  | Small |  | 1043 |
|  | Very Small |  | 983 |
| Commodity Sampled | Chicken | Raw intact | 206 |
|  |  | Raw ground, comminuted or nonintact | 392 |
|  |  | Young carcass rinse | 520 |
|  | Turkey | Carcass sponge | 43 |
|  |  | Raw ground, comminuted or nonintact | 53 |
|  | Beef | Raw intact | 1180 |
|  |  | Raw ground, comminuted or nonintact | 1377 |
|  | Pork | Raw intact | 157 |
|  |  | Raw ground, comminuted or nonintact | 443 |

Our analyses use a custom fork of the GNNWR Python package developed and described in Yin et al. (2024). In particular, we modified the GNNWR package’s codebase to exponentially transform the predicted response value — constrained at 20 to prevent computational overflow and to avoid evaluating predicted responses antibiotic resistant samples larger than *e*^20^ during the model training stage — and allow using a Poisson NLL loss function to evaluate the overall regression. Additional custom R (R Development Core Team, 2008) and Python (Python Software Foundation, 2025) scripts were developed for collating the FSIS data and carrying out the analyses and visualizations. All code and data used in the analyses, including hyperparameters for fitting the pGTNNWR, are released under the GNU Public License v3 (Stallman, 2007) and are publicly available at https://github.com/kewok/amr_meatplants.

## Results

To facilitate interpretation, we provide visualized results in the main text from five, evenly spaced seasons during the time horizon analyzed: Autumn 2021 (2021.4), Summer 2022 (2022.3), Spring 2023 (2023.2), Winter 2024 (2024.1) and Autumn 2024 (2024.4). Results for the entire time horizon analyzed are provided in Supplementary Material S1. The seasonal trends characterized there are broadly akin to those we present in the main text. The overall Residual Mean Standard Error (RMSE) of the resultant pGTNNWR was 0.916. While there appeared to be consistency in summary statistics over sufficient chain/epoch lengths, we did observe some transient variability in measures of model performance during the initialization stages (up to about timestep/epoch 50-100 out of several thousand epochs) over differing random seeds and batch sizes.

Figure 2A-E describes the number of predicted AbR positive samples at each plant in our dataset. Fig. 2F-J shows the natural logarithm of the absolute residual. The pGTNNWR suggests more cases of AbR resistance to be found along the US Atlantic seaboard than in other regions, although in some seasons predicted regional differences were not as pronounced (Fig. 2A-E). However, the pGTNNWR appeared to be somewhat more error-prone particularly along the Atlantic coastal regions in the Middle Atlantic — ranging from northern Florida and Georgia in the South to southern New England in the North (Fig. 2F-J). In earlier seasons (Fall 2021—Fall 2022), the model displayed less predictive accuracy in the southern Mississippi Valley than later seasons. In general, the pGTNNWR appeared most consistently accurate in the eastern Great Lakes region (Fig. 2F-J). Although there were sporadic, strong overpredictions starting in Fall 2022, the model otherwise appeared to generally perform comparatively across seasons (Fig. 2F-J).

**Figure 2.**
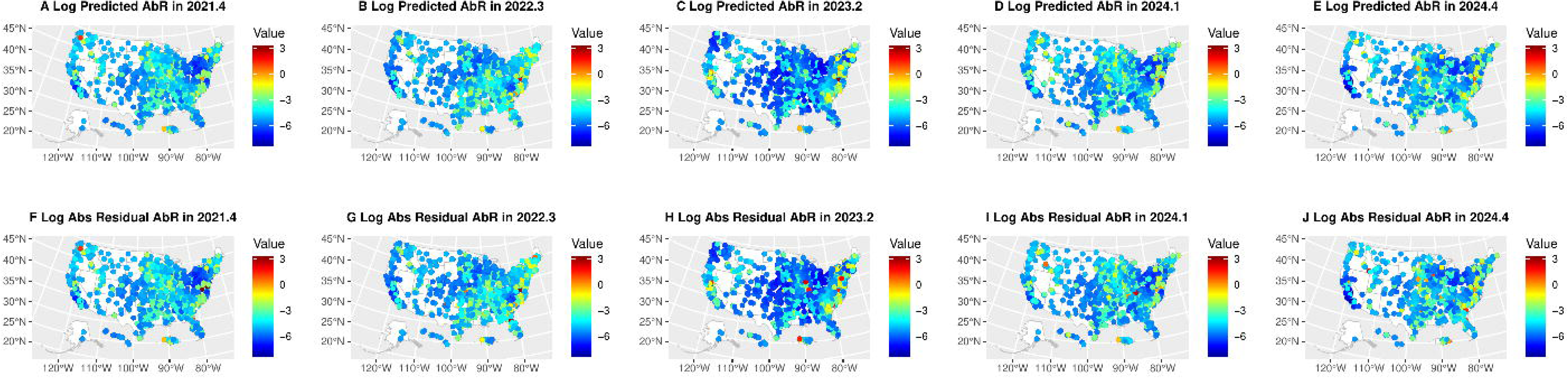
The log number of antibiotic resistant (AbR) positive samples predicted by the pGTNNWR in each meatpacking facility across the United States, as well as the log absolute residuals during these same time periods. Here, and in subsequent figures, panels in the leftmost column depict results for Fall 2021, panels in the second-to-left column Summer 2022, panels in the central column Spring 2023, panels in the second-to-right column Winter 2024 and panels in the right-most column Fall 2024

Figures 3–6 describe the effect of each of our predictors on the number of predicted AbR positive samples across space and time. Below, we characterize the seasonal and regional heterogeneities we found in how each of these predictors affect the number of positive AbR samples for each plant.

**Figure 3.**
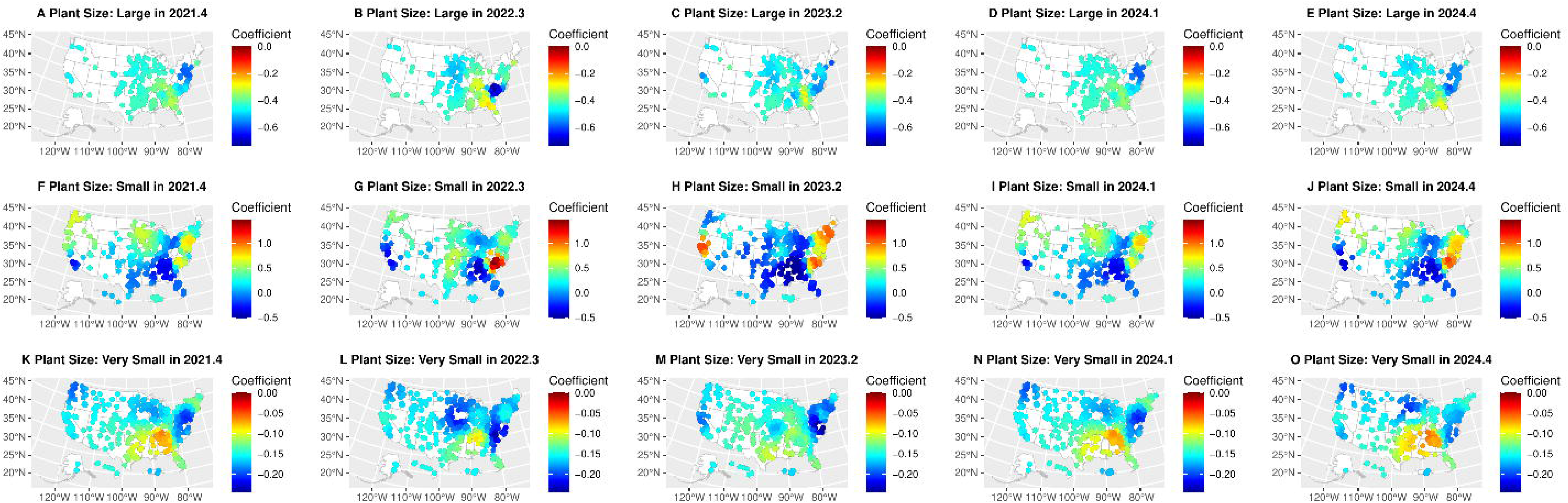
Spatiotemporally weighted regression coefficients for the effect of plant size for (A-E) large, (F-J) small and (K-O) very small facilities on the number of AbR positive samples across seasons throughout the United States.

**Figure 4.**
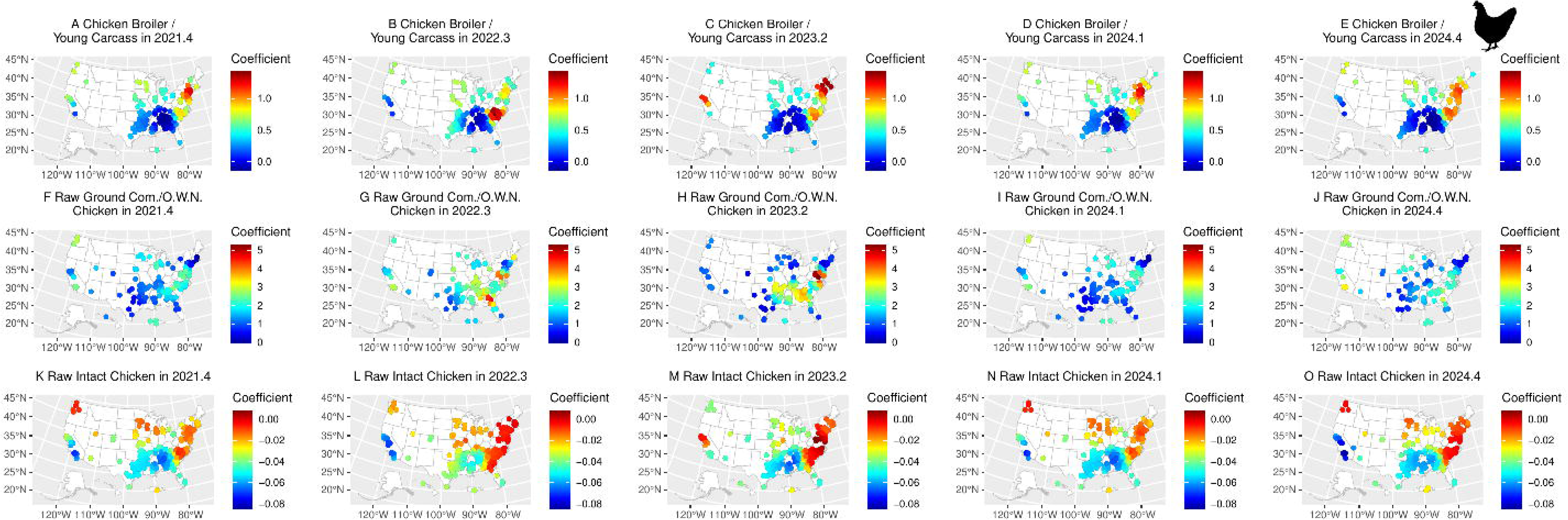
Spatiotemporally weighted regression coefficients for the effect of handling (A-E) chicken broilers and chicken carcasses, (F-J) raw ground, comminuted and otherwise nonintact (Com./O.W.N.) chicken and (K-O) raw intact chicken on the number of AbR positive samples across seasons throughout the United States. Credit: Arcadia Science 2025 CC0 1.0 Universal.

**Figure 5.**
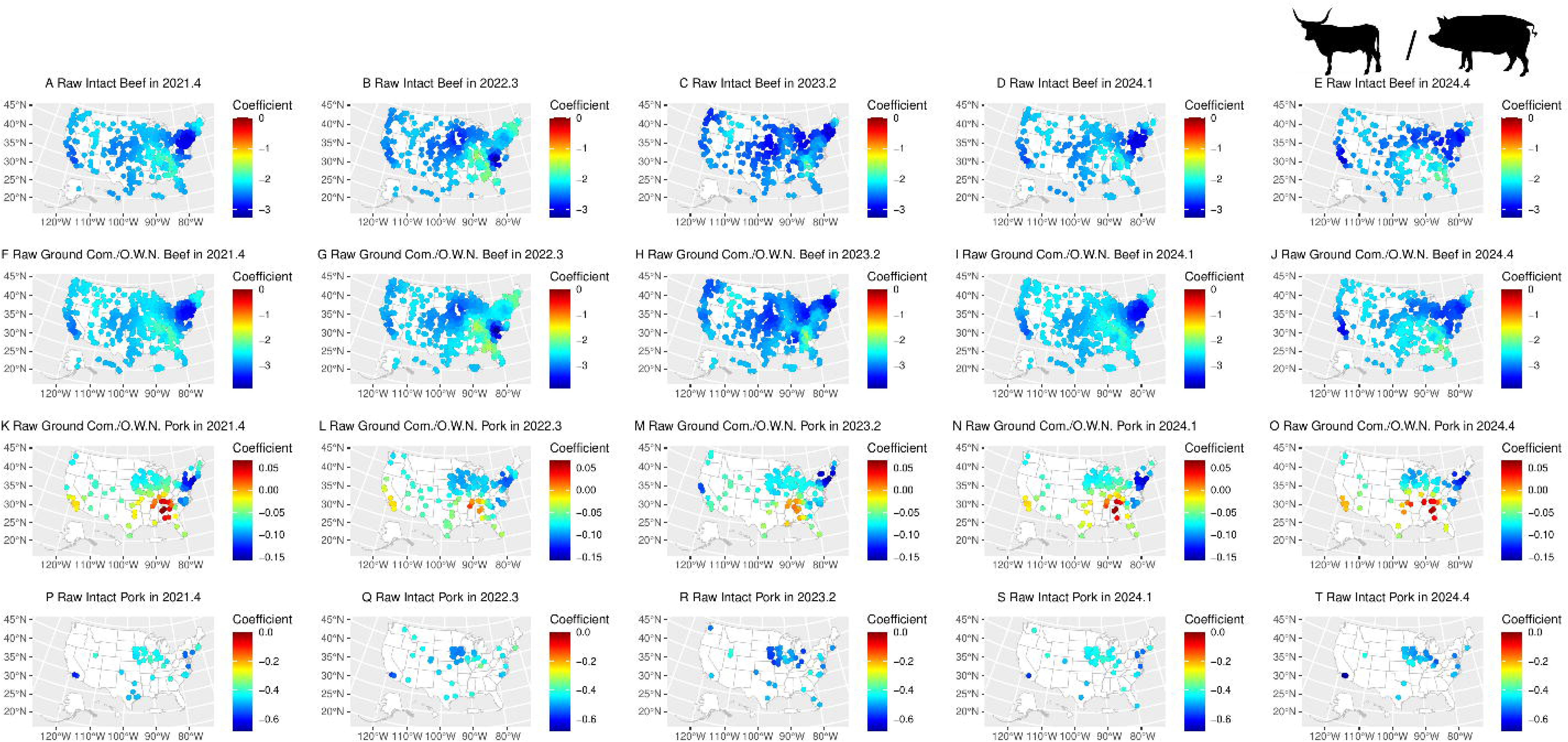
Spatiotemporally weighted regression coefficients for the effect of handling (A-E) raw intact beef, (F-J) raw ground, comminuted and otherwise nonintact (Com./O.W.N.) beef, (K-O) raw ground, comminuted and otherwise nonintact pork, and (P-T) raw, intact pork on the number of AbR positive samples across seasons throughout the United States. Credit: Tracy Heath 2013 CC0 1.0 Universal and Steven Traver 2012 CC0 1.0 Universal.

**Figure 6.**
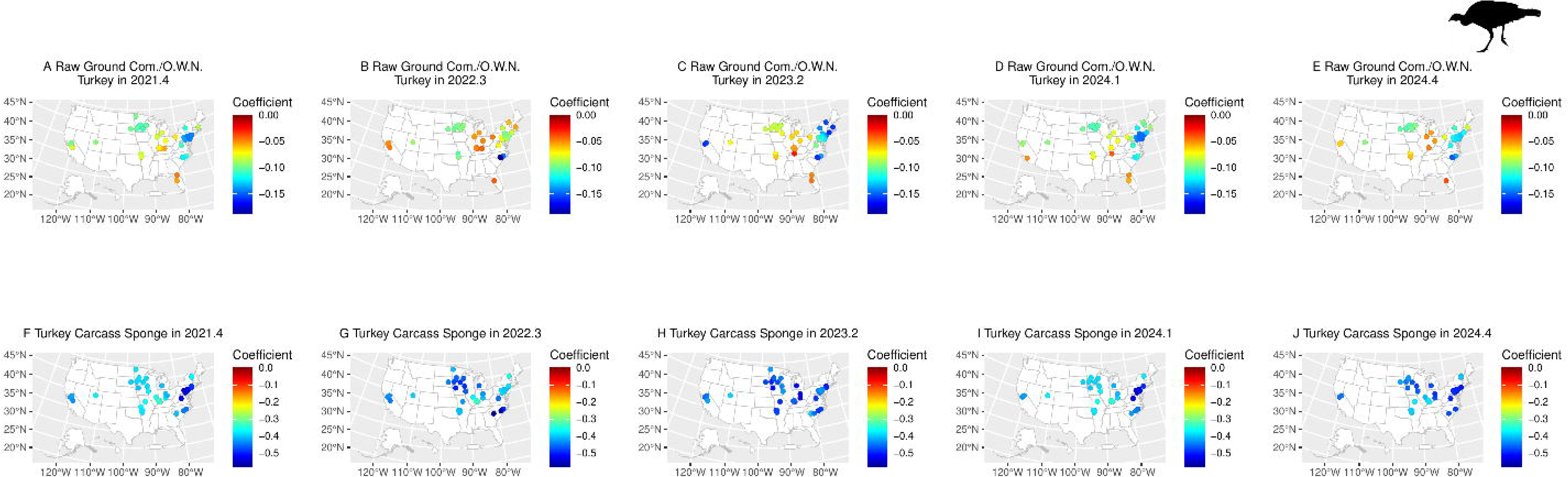
Spatiotemporally weighted regression coefficients for the effect of handling (A-E) raw ground, comminuted and otherwise nonintact turkeys, (K-O) turkey carcasses and sponges on the number of AbR positive samples across seasons throughout the United States. Credit: Abby Weber 2025 CC0 1.0 Universal.

Perhaps unsurprisingly, our results suggest plant size has a sharp impact on AbR positivity (Table 2s). However, somewhat unexpectedly, the largest effects on the number of AbR positive samples tend to come not from larger plants, where there would presumably be a greater diversity of product sources (Fig. 3A-E), but plants between the extrema of large and very small sized facilities (designated as “small” in the dataset) (Fig. 3F-O). Being such a plant along both the Pacific and Atlantic Coasts appeared especially likely to contribute to the number of AbR positive samples. This is in stark contrast to the effect of being a small plant in two other regions: (i) the Midwest / Mississippi Valley and Rocky Mountain West. In the Midwest / Mississippi Valley, small plants appeared to contribute less to the number of AbR samples detected, whilst being a very small plant in the southern range of these regions had an amplifying effect on AbR positivity. By contrast, in the Rocky Mountain West, meatpacking plant size appeared to have relatively middling effect sizes (relative to the overall national range) on the number of AbR positive samples across facility sizes. Along the Pacific Coast, there was also some indication of a North-South cline in terms of how much effect being small or very small plants had on the number of AbR samples. Being a very small plant in the Pacific Northwest, for instance, could reduce the number of AbR positive samples relative to very small plants in the Southwest (particularly in Fall 2021 and throughout 2024). By contrast, among small plants the trend tended to be reversed. On the Atlantic Coast, plant size had especially varying effects in the Mid-Atlantic region. There, being a small plant elevated the number predicted AbR positive samples relative to very small and large plants.

Figure 3 also illustrates temporal differences in the regional effects of plant size. The effects of being a small plant appear strongest, for example, between Winter 2022 and Summer 2023 in the southern part of the US Atlantic Coast north of Florida (Fig. 3F-J). Similarly, the effects of being a very small plant (Fig 3K-O) or a large plant (Fig 3A-E) in the Southeast shift from being comparatively smaller than other regions in the middle of the time horizon we considered to being more pronounced than other regions later in the time period.

Assessing the effects of plant size on AbR positivity suggests even for predictors with an especially strong effect on AbR positivity, these effects vary substantially across space and seasons. Distinct spatial effects of plant characteristics on AbR positivity also become apparent when chicken products are considered (Fig. 4). A key result is that the nature of regional heterogeneities in how being a chicken-handling meat processing plant contributes to AbR positivity differs noticeably by the kind of chicken product handled. For instance, plants handling chicken broilers and young chicken carcasses and raw intact chickens on the Atlantic Coast appear to elevate the number of AbR positive samples relative to such plants in the Lower Mississippi Valley and inland Southeast (Fig. 4A-E and 4K-O). By contrast, handling raw ground, comminuted, and otherwise nonintact chicken had less effect in these regions (Fig. 4F-J). Such varying regional effects between different chicken products can also be seen in the Upper Midwest and Northeast US. There also appeared to be some seasonal effects in regions of the Pacific Coast in the handling of broilers and raw intact chickens, although the temporal differences in these effects are less stark in the Pacific Northwest (Fig. 4A-E and 4K-O). In other regions, subtle temporal trends were also observed. For example, the effect of handling non-intact chicken on AbR positive samples increases in late 2022 and early 2023 in the Southeast and southern New England relative to other seasons examined.

Regional differences in the effects of facilities handling beef and raw intact pork (Fig. 5A-J and P-T) appear somewhat milder than the strong regional signatures of the effects of differing plant sizes and chicken-derived commodities on AbR described above. To be sure, we find a general trend of increasing importance of handling beef products on AbR positive samples moving from the US Pacific Coast and Rocky Mountain regions to the southern Atlantic Coast (Fig. 5A-J). Some seasonal effects, particularly in the Pacific Northwest, the Upper Midwest, and New England can also be seen for both beef commodities. A similar, if perhaps subtler, temporal gradient also becomes apparent for the effect of handling raw intact pork on AbR positivity (Fig. 5P-T). Despite these spatiotemporal heterogeneities, the regional and seasonal differences on whether a plant handles these commodities do not appear as pronounced as was the case for facility size and certain chicken commodities discussed above. We add, however, that raw ground/non-intact pork appears to have spatial signatures somewhere in between the relatively muted effects of handling beef products and intact pork, on the one hand, and the notable variability in the effects of plant size and handling chicken products, on the other (Fig. 5K-N). There is generally a tendency towards decreasing importance of processing ground/nonintact pork in the Northwest and Northeast, as well as a North-South cline throughout the US. Thus, viz. the number of AbR positive samples, the effects of a facility handling raw ground and nonintact pork (Fig. 5K-N) more closely resembles the effects of a facility for raw ground and nonintact chicken (Fig. 4F-J) across space than the region-specific effects of a facility handling raw intact pork.

Finally, Figure 6 shows regional trends in the effect of processing turkey meat on AbR positivity differed depending on the commodity in question. Where a plant was located appeared to impact how strongly processing raw ground and non-intact turkeys affected the number of AbR positive samples (Fig. 6A-E), with the effect stronger for facilities in the Western US, especially from Autumn 2021 through Summer 2022 (Fig. 6A, B). Yet spatial heterogeneity was not as pronounced on the effect of handling turkey carcass sponges, although there was some indication of temporal variability in this effect in the Upper Mississippi Valley, with the effect of handling turkey carcass sponges there being larger than elsewhere early and late in the time horizon we analyzed (Fig. 6F-J).

## Discussion

Despite their potentially critical role in facilitating and mitigating the spread of antibiotic resistant (AbR) bacteria, few studies have aimed to capture both how and why the risk profiles of differing meat packing operations shift across large spatiotemporal arenas. In this study, we presented and applied a novel method — Poisson geographic and temporal neural network weighted regression (pGTNNWR) — that extends the use of neural network-based spatiotemporal regression modeling to analyze the number of samples positive for antibiotic resistance in meatpacking facilities. Our pGTNNWR regressed predictor variables — plant size and commodity type — on the number of AbR positive samples collected at each site during a given season.

While variegated, our results convey a core theme: space and time play a foundational role in the appearance of AbR in US meat supply chains, however expansive, just-in-time, and, upon processing, refrigerated the latter may be (Lin et al., 2019; Mulik, 2024; Miller et al, 2025). On some level, this may seem unsurprising. In an economic ecology as diverse as a production sector across major food commodities, operations, and seasons on what is essentially a continental scale, not all meatpacking facilities can be expected to exhibit equally likely potential for AbR positivity if by chance alone. But our study lends credence to the notion that the epidemiological consequences of key explanatory factors we consider — in particular, plant size and commodity handled — are contingent on specific sectoral, regional, and seasonal contexts.

To be sure, there are undoubtedly a large number of other salient features of meat processing facilities than those we considered here that may explain patterns of AbR positivity. To name just a few, the diversity of farms from which animals are sourced, a plant’s profit margins, whether the processing plant is an independent operation or part of a larger corporate network, the extent of vertical integration for different commodity types, and whether the processor sells directly to consumers could all potentially impact the number of AbR positive samples (Wallace & Kock, 2012; Crespi & Saitone, 2018; Lin et al., 2019; Miller et al., 2025). For instance, does the greater AbR exposure for medium-sized processing plants found here represent a convex diseconomy of scale based in limited capitalization and overhead for microbial testing (Ollinger et al., 2005; Ollinger & Moore, 2009; Ollinger & Bovay, 2018)? Is AbR seasonality to be found in bacterial ecologies operating outside the bounds of model expectations of continuous livestock and poultry production (Steuernagel et al. 2023; Farkas et al., 2025; Kangogo et al. 2025)?

Our results suggest a more ambitious examination integrating these additional drivers might provide further insight into what explains spatiotemporal heterogeneity in AbR across meat packing facilities. Nevertheless, such an approach faces constraints common to all research projects, not least of which is data availability. Ultimately, there are only so many meat processing plants in the US, and with factorial (e.g., commodity type), rather than quantitative, predictors, much more extensive collection records would be needed to permit reliable quantitative analyses of the sort we have performed here. One potential way forward may be to narrowly target a more comprehensive analysis for a given commodity category — e.g., raw intact beef — and identify ways in which additional predictors could be systematically quantified along one or a few axes (such as using information-theoretic measures to characterize supplier diversity —as reviewed in Sivadasan et al., 2006; Isik, 2010; Cheng et al., 2014). At present, all our predictor variables are discrete factors, rather than continuous predictors. Thus, each plant size and commodity becomes its own indicator variable (e.g., each facility is scored as either “0” or “1” for large plant size, “0” or “1” for small plant size, and so on), which ultimately increases the sparseness of the dataset on which the model trains.

Although an overall RMSE of 0.916 may seem high, an examination of run-time statistics showed repeated randomizations of the training, testing, and validation data do not improve the pGTNNWR’s negative log likelihood score after several hundred epochs (Supplementary Material S2). This suggests that despite the considerable extent of the FSIS records, data availability may still constrain the model’s overall performance. The explanatory capacity of a modeling strategy such as ours may thus be strengthened if continuous, quantitative predictors characterizing plants (e.g., number of employees, profitability or number of farms from which the animals come) can be identified and be associated with each plant. This will allow pGTNNWR to be fitted along a more fine-grained set of input values.

A further methodological point we highlight is that while inspired by Poisson Deep Neural Networks (PDNNs) (e.g., Montesinos-López et al., 2020; Montesinos-Lopez et al., 2021), strictly speaking, pGTNNWR is not precisely analogous to a PDNN. This is because PDNNs directly use the output layer of the neural network itself to predict the response values. By contrast, in our approach, the neural network is constructed in two stages: first, to characterize the distancing metric in space and time (Fig. 1A(iii)), and second, to infer the spatiotemporal weights for the coefficients *β_i_*(·) of the regression equations (Fig. 1A(iv)). The output of these neural networks is then further filtered through a linear combination among factors and their weighted coefficients to generate predicted response values. To our knowledge, our study is the first to apply the basic idea of pGTWNNR — whereby the predicted response is exponentially transformed and the model evaluated with a Poisson NLL loss function — to a geographic and temporally weighted regression context. As such, this approach could potentially also apply to other epidemiological count data, which often appear when examining geographic and temporal heterogeneities in disease incidence. We therefore propose our study provides a useful advance to understanding the role of potentially spatiotemporally heterogeneous processes in driving disease dynamics.

We further suggest that our technique is uniquely well-suited for epidemiological studies of emerging infectious diseases such as AbR bacteria. Strategies such as the one we propose can shed novel insights on the downstream evolutionary epidemiological consequences of broad-scale socioeconomic trends (Aguilar-Støen et al., 2025). Of note, in recent decades there has been a marked consolidation of meatpacking operations in the United States (reviewed in MacDonald et al., 1999; Jakobsen & Hansen, 2020; McKendree et al., 2021; Saitone et al., 2023; Rowe, 2024). This consolidation, in turn, has likely altered bacterial ecosystems over space and time, with implications for the management of AbR (Jakobsen et al., 2025). For instance, by 2022 the four largest broiler processing firms accounted for 57% of US operations, with survey data indicating 50% of contract poultry growers had access to only one or two live poultry dealers (MacDonald & Key, 2012; Aparicio et al., 2021; WATT PoultryUSA, 2021). These developments reflect a production system in which integrators increasingly supply chicks to growers who are contractually required to raise the birds and sell them back to processing plants operated directly or indirectly by those same firms. (Wallace & Kock, 2012; McKenna, 2017; USDA, 2023). This has entailed rerouting the movement of chicken hosts and, likely inevitably, their bacterial (as well as viral and parasitic) symbionts through reconfigured distribution networks over both space (Fournié et al., 2013; Guinat et al., 2023) and time. Transit and processing schedules are now highly syncopated (Thompson & Applegate, 2008; Adamczuk Oliveira & Lindau, 2012; Cobb, 2020; Kwon et al., 2021). By assessing when, where, and how AbR bacteria are detected at the plant level, we can begin to elucidate how continent-scale industrial shifts, such as the adoption of greater degrees of vertical integration for chicken meat production, can impact epidemiological outcomes by altering host-bacteria ecosystem dynamics, and hence AbR selection pressures, over space and time.

The potential implications of the long-term consolidation of the US meatpacking industry for disease control are not just confined to the effectively continental landscape we examined over epidemiologically relevant time horizons. Such sectoral changes can also alter the public health impacts of AbR bacteria at the individual plant level. For instance, several studies have documented increased effluence at individual meatpacking plants, including plants operated or otherwise controlled by large scale, multinational meat processing companies (La Rosa et al., 2025; Xu et al., 2025). The extent to which AbR bacteria and/or their genetic material — plasmids or otherwise — potentially present in this sewage is highly variable and its impacts on human AbR infection incidence remain poorly understood (Bengtsson-Palme et al., 2018; Savin et al., 2021; Chau et al., 2022; Foyle et al., 2023; Toribio-Celestino and Millan 2025). Yet by illustrating when and where different kinds of meat processing plants may be associated with AbR detection probabilities, this study and others can begin to provide an open-source resource for communities in which processing plants are located or proposed to be built to systematically assess the risk profiles local operations may or may not present viz. AbR bacteria.

Public disclosure and even shaming have had their demonstrated impact upon plant-to-plant food-safety mitigation (Ollinger & Bovay, 2020; Bovay, 2025). Finer operationalization in public health policy is also possible. For instance, institutional cognition — governmental or community-based — can act on the knowledge of whether specific genotypes of human cases of AbR-related infection as detected in area hospitals are related to those tested for in local processing plants (Mulvey et al. 2005; Crippa et al., 2025; Pereira et al., 2026). The USDA FSIS data are parsed by bacterial lineage. We might ask whether some plants support specific strains or greater diversities in bacterial species/serotypes by plant size, commodity, and/or farm catchment (Asmus, 2025; Brito et al., 2025).

Finally, in many parts of the US, the worker compositions of meatpacking plants have shifted markedly to reflect declining union membership, targeted hiring of newly arrived immigrants, higher turnover rates and increased worker precarity (McConnell, 2019; Luckstead et al., 2022; Fukushima et al., 2024). Moreover, practices across plants, including the adoption of novel techniques for blood and meat handling, are increasingly becoming standardized over space (USDA FSIS, 1996, 2021a). The shift towards large corporate ownership of individual facilities has been hypothesized to have accelerated each of these within-plant trends (MacDonald et al., 1999; iPES FOOD, 2017; Green, 2020; Barcenilla et al., 2024). We therefore suggest studies such as ours have critical implications for occupational health and safety (see also Ruiz et al., 2025). As sectoral trends alter workforce composition and labor conditions, as well as the characteristics of surrounding communities from which meatpacking plant workers come, understanding the spatiotemporal dynamics of AbR exposure risks faced by plant workers and the communities in which they live will be essential to conducting effective public health interventions (Woteki & Kineman, 2003; Su et al., 2019; Freshour, 2019; Crippa et al., 2025).

To conclude, our proposed strategy investigating the distribution of AbR detection patterns at intermediate and highly interlinked nodes of the meat supply chain can provide critical first steps towards elucidating whether and how selection for AbR at and beyond the level of animal husbandry operations cascades to the increased human AbR infectious disease burden. Especially within the context of a major production sector (such as meat processing) rapidly in flux, identifying how the effects of drivers of AbR positivity vary over space and time can provide distinct perspectives on the targeted, regionalized, and localized disease control and management strategies. Our approach provides a framework that could be potentially applicable for other diseases and thinking about the dynamics and spread of emerging infectious pathogens over broad spatial scales.

## Data Availability

All data produced are available online at https://github.com/kewok/amr_meatplants

## Acknowledgments

The authors would like to thank Patrick Kerrigan (1959-2021) and Jay Shooster for insightful conversations early in the project, as well as for helping us better understand the structure and organization of the database upon which this study is based. HGP received support from the College of Arts and Sciences and the Undergraduate Research Opportunities Program at the University of St. Thomas. No other funding was received for this project.

## Supplementary Material

S1. (a) Log model predictions for every season in the dataset and (b) the log absolute residuals. The spatiotemporally weighted regression coefficients for every season for the effects of being a (c) large, (d) small or (e) very small facility. The spatiotemporally weighted regression coefficients for each season for the effects of being a facility handling (f) boilers and young chicken animals, (g) ground, comminuted or otherwise nonintact chicken, (h) raw intact chicken, (i) raw intact beef, (j) ground, comminuted or otherwise nonintact beef, (k) ground, comminuted or otherwise nonintact pork, (l) raw intact pork, (m) ground, comminuted or otherwise nonintact turkeys and (n) turkey carcasses and sponges.

S2. Loss function evaluation for the training (solid) and validation (dashed) data partitions during model fitting.

