## Supplementary figures and images for "Seasonal and regional heterogeneities in antibiotic resistance (AbR) across meatpacking plants in the USA"

### S1a.pdf-1.png

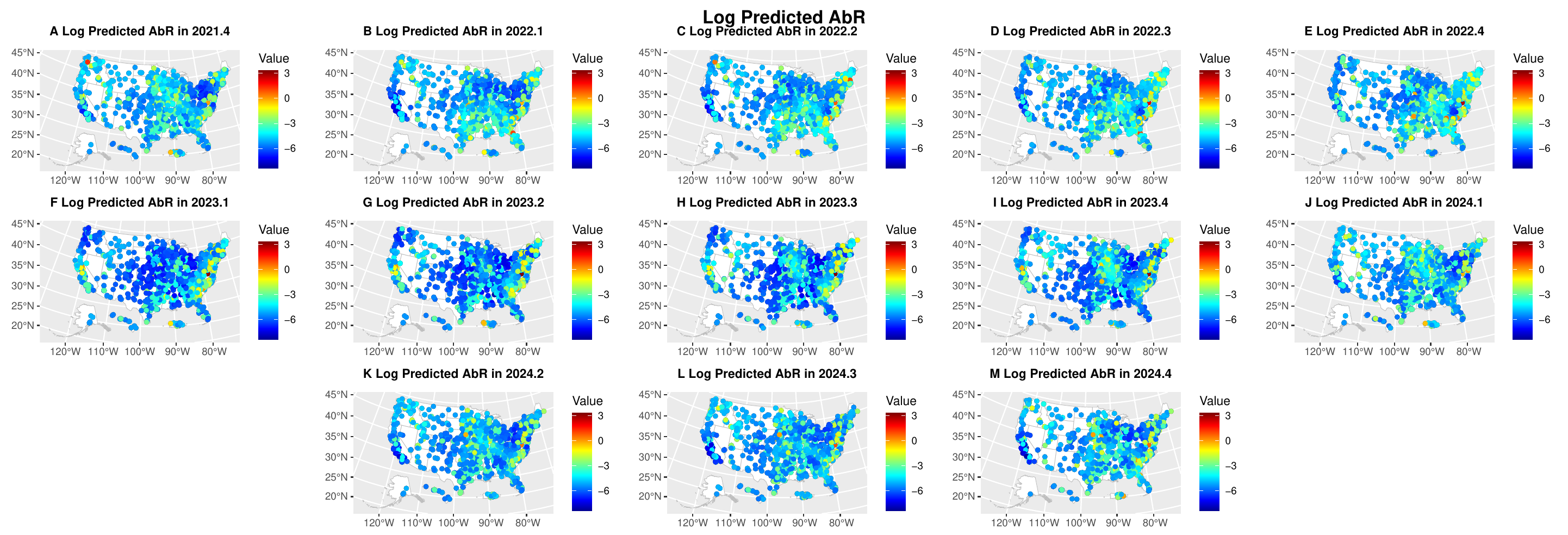

### S1b.pdf-1.png

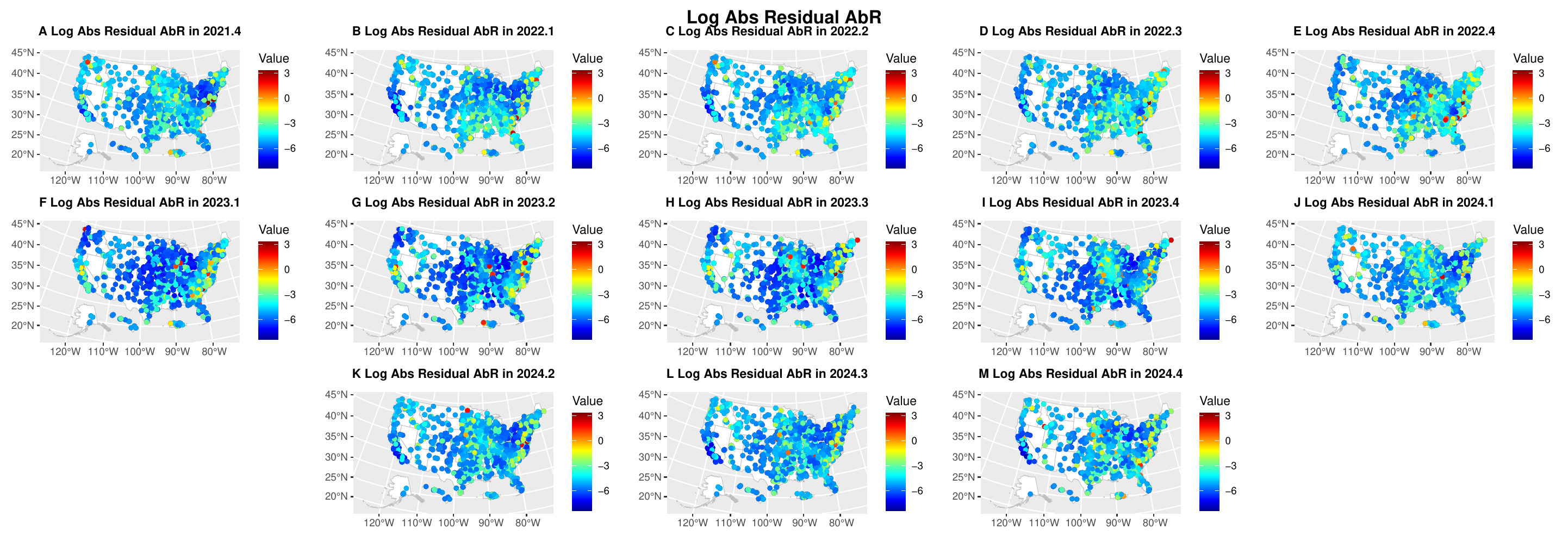

### S1c.pdf-1.png

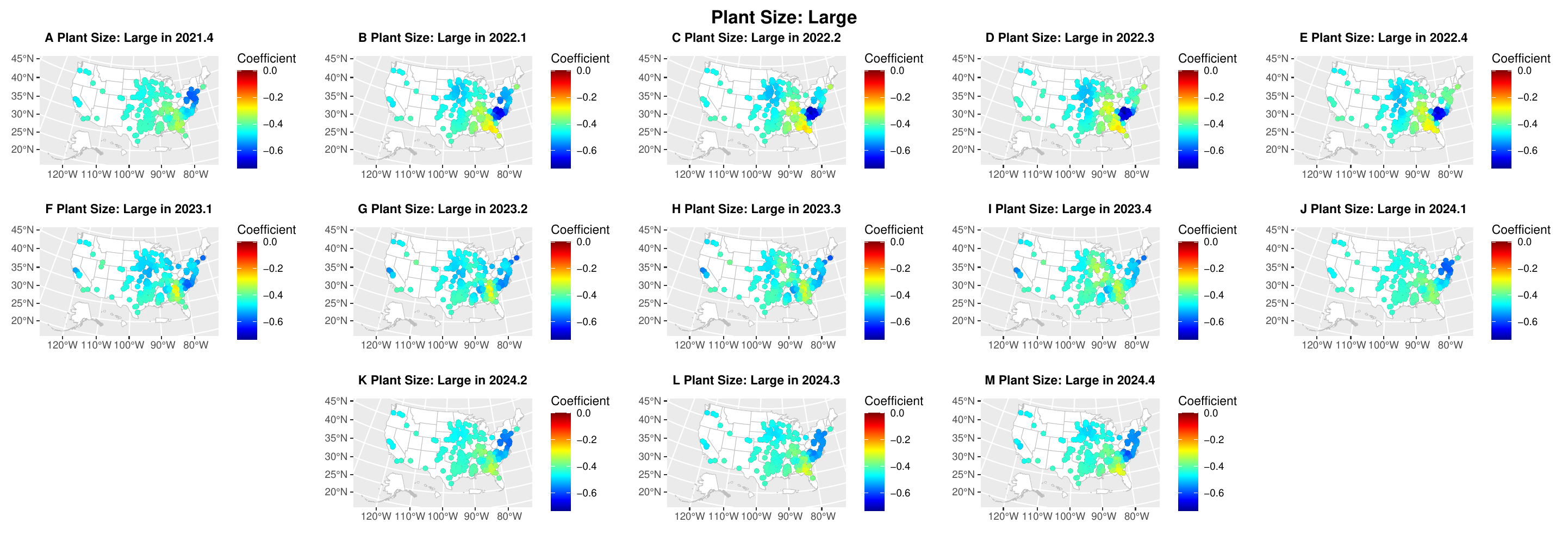

### S1d.pdf-1.png

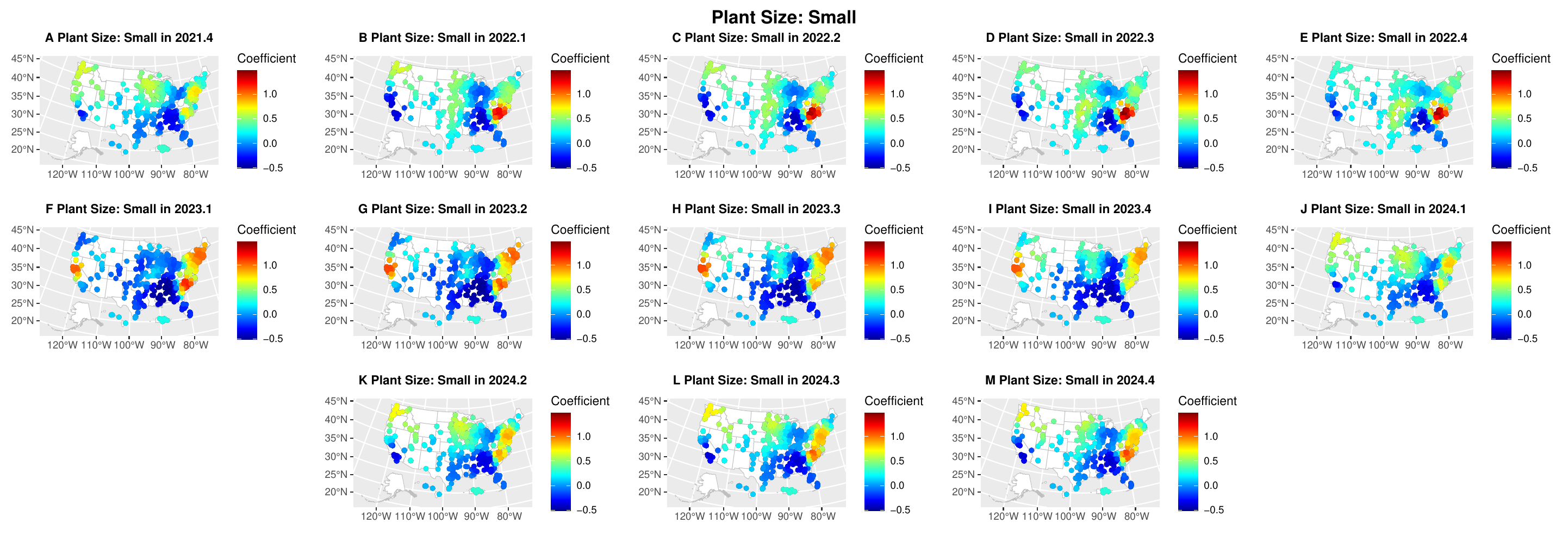

### S1e.pdf-1.png

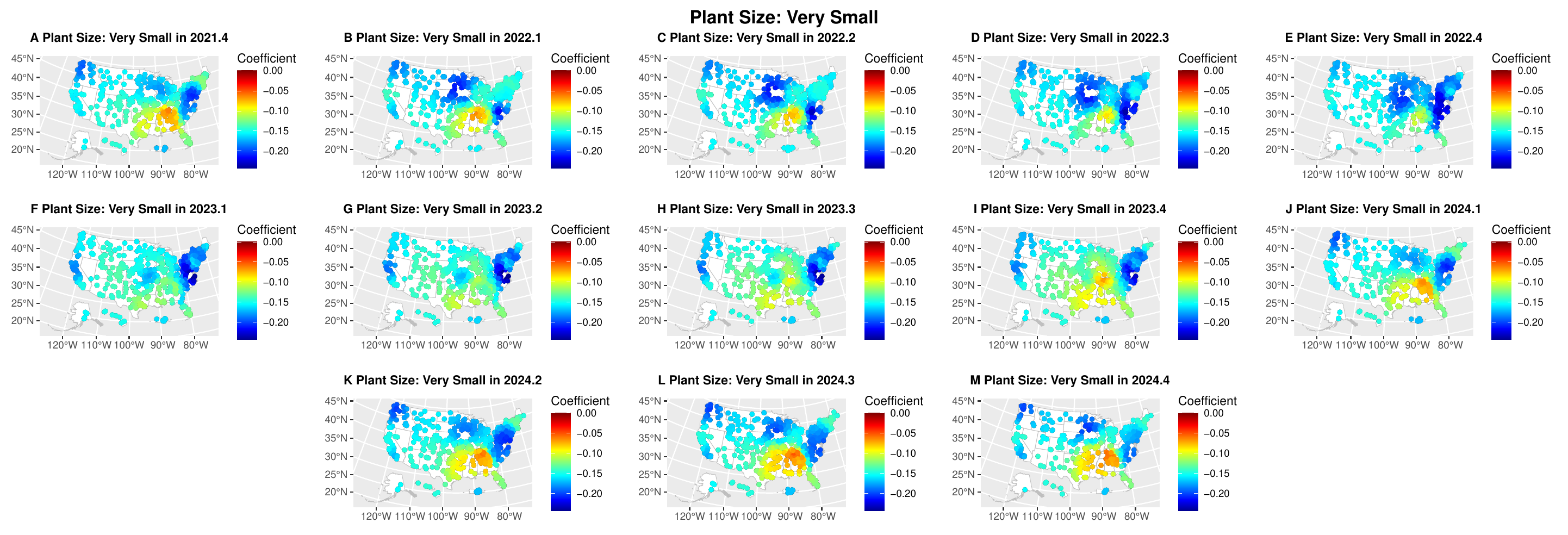

### S1f.pdf-1.png

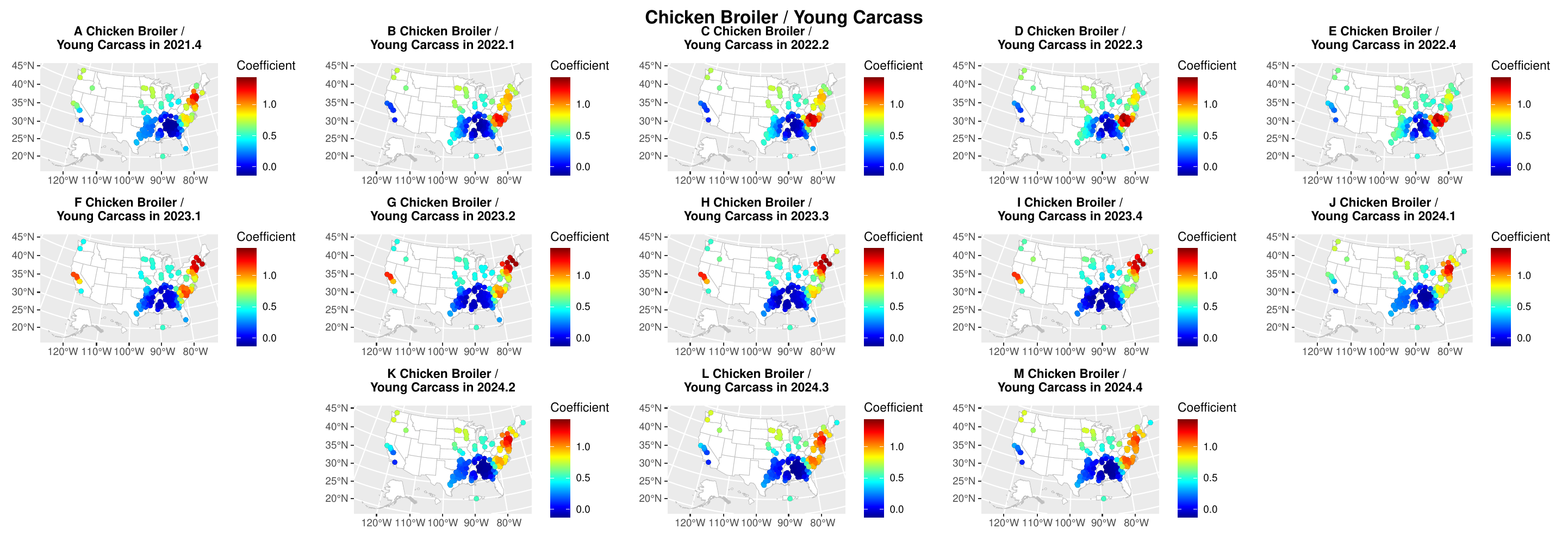

### S1g.pdf-1.png

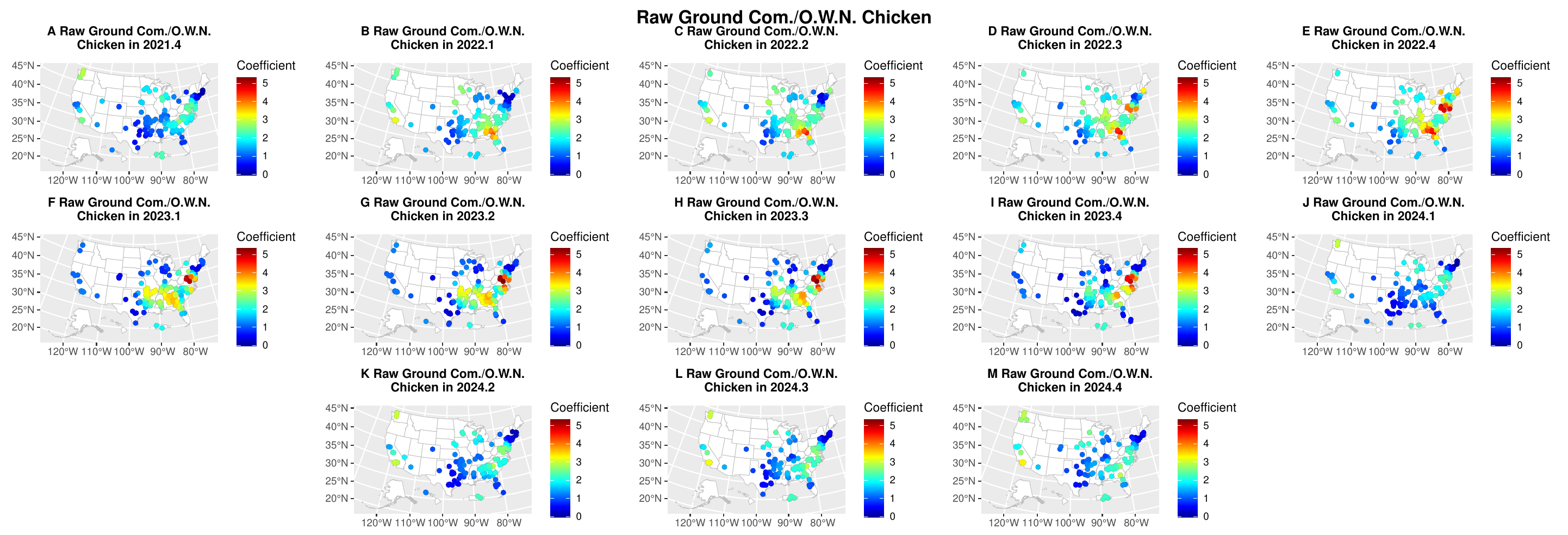

### S1h.pdf-1.png

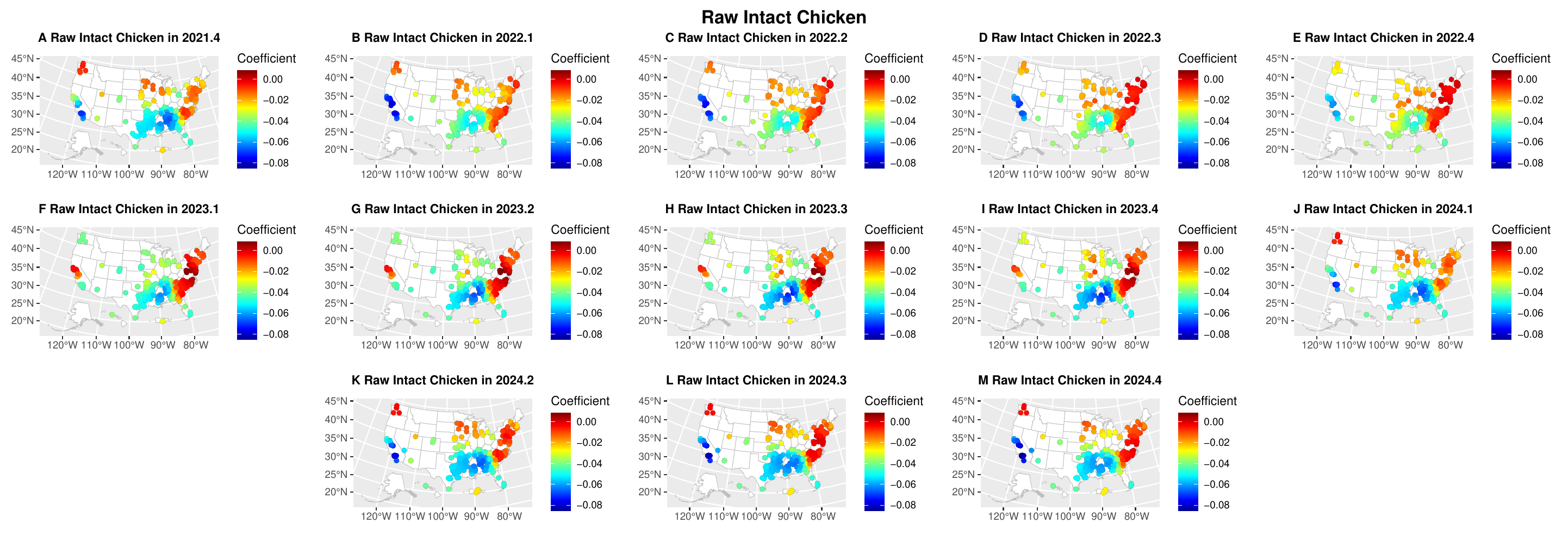

### S1i.pdf-1.png

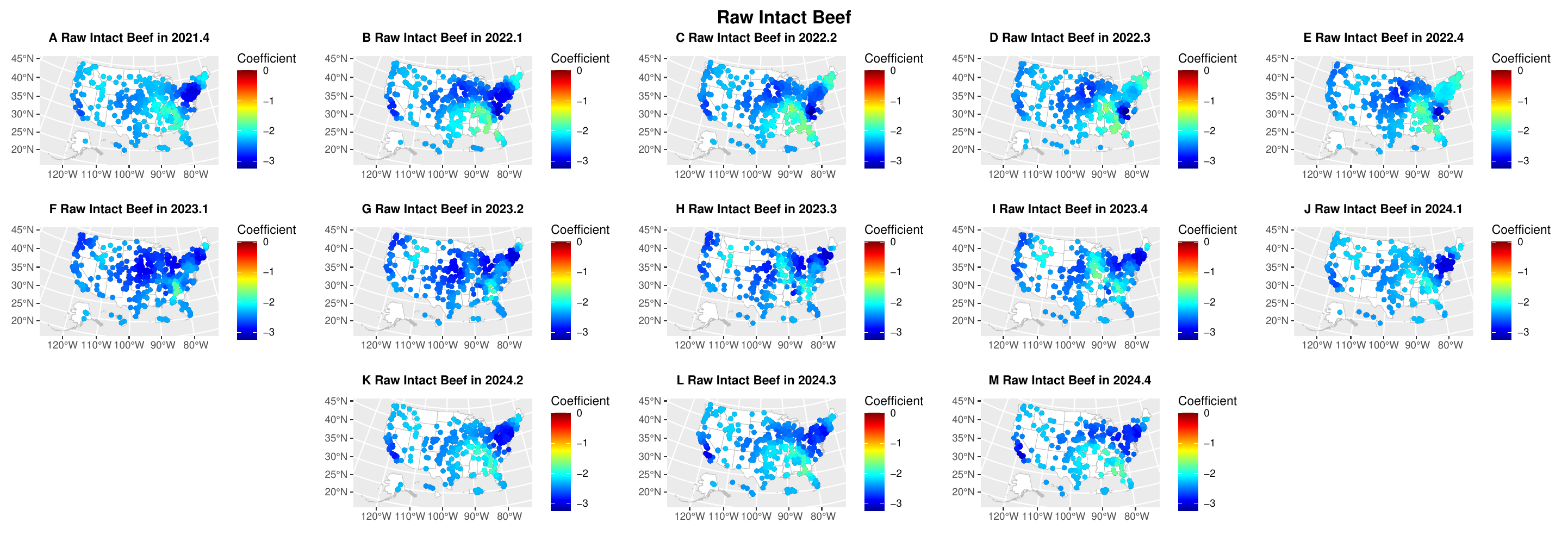

### S1j.pdf-1.png

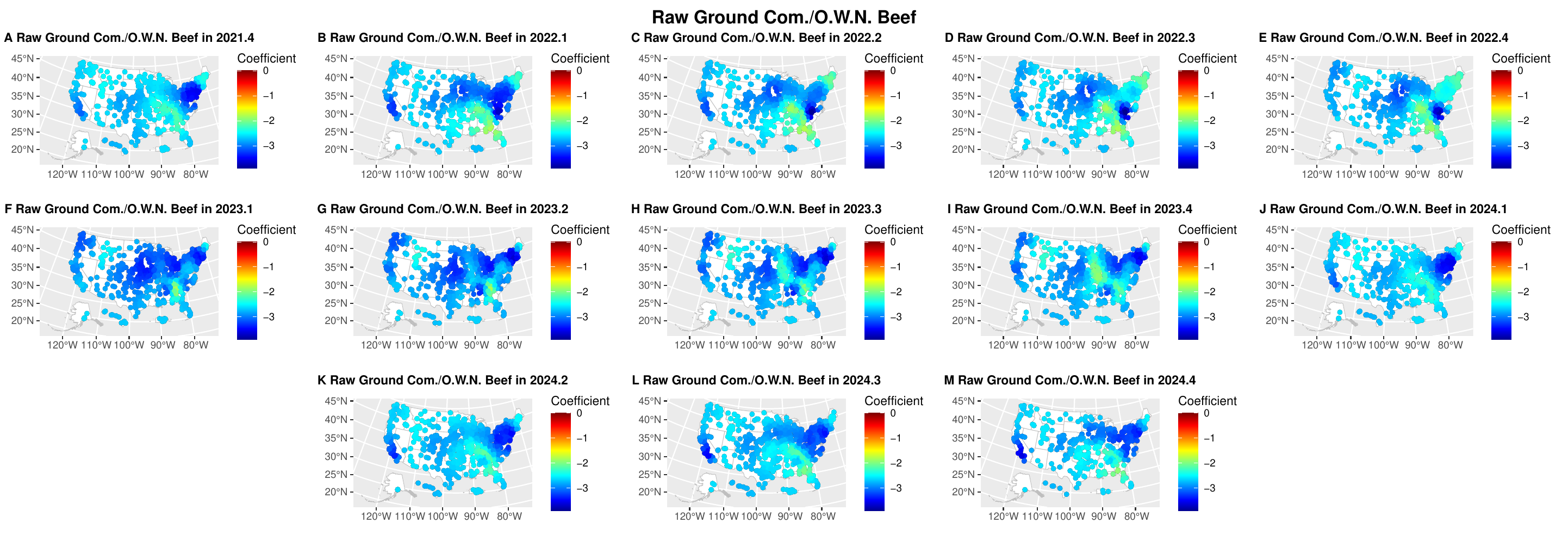

### S1k.pdf-1.png

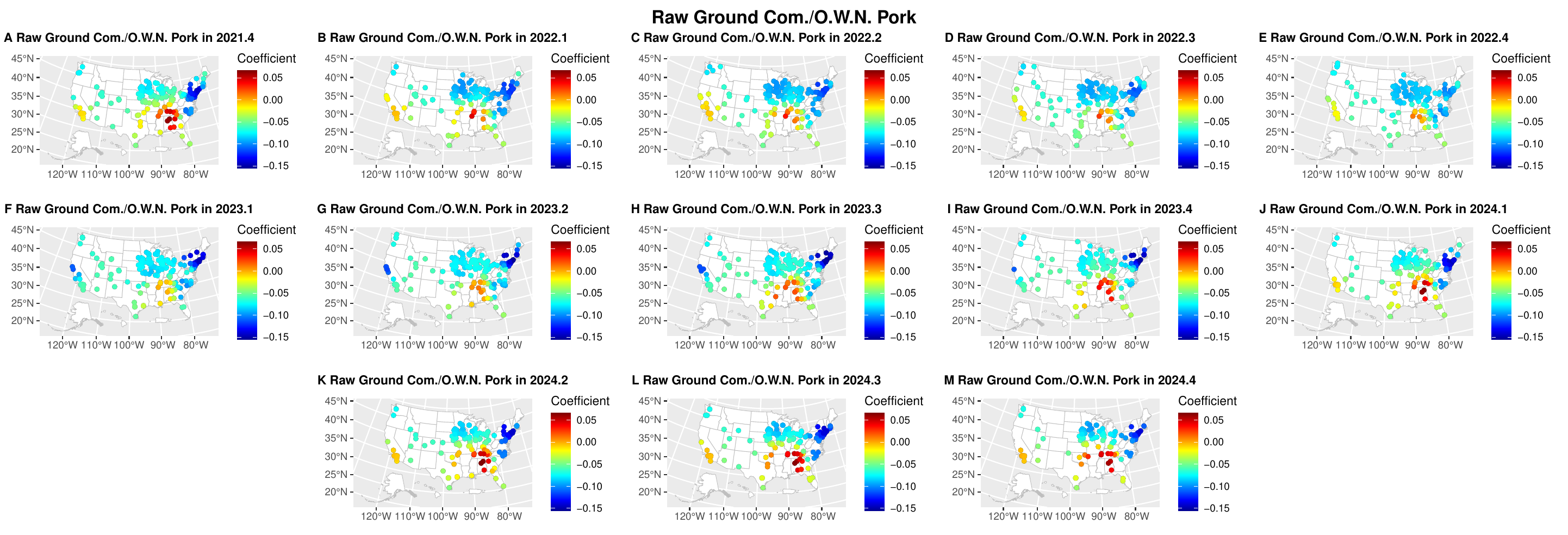

### S1l.pdf-1.png

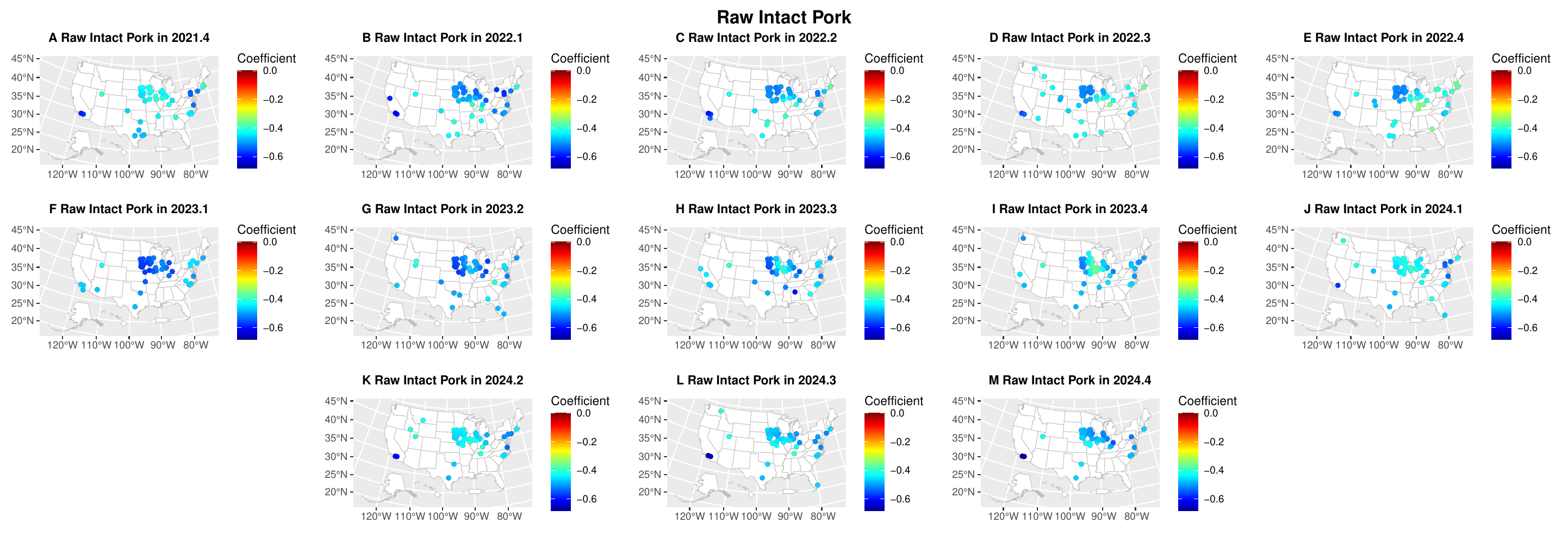

### S1m.pdf-1.png

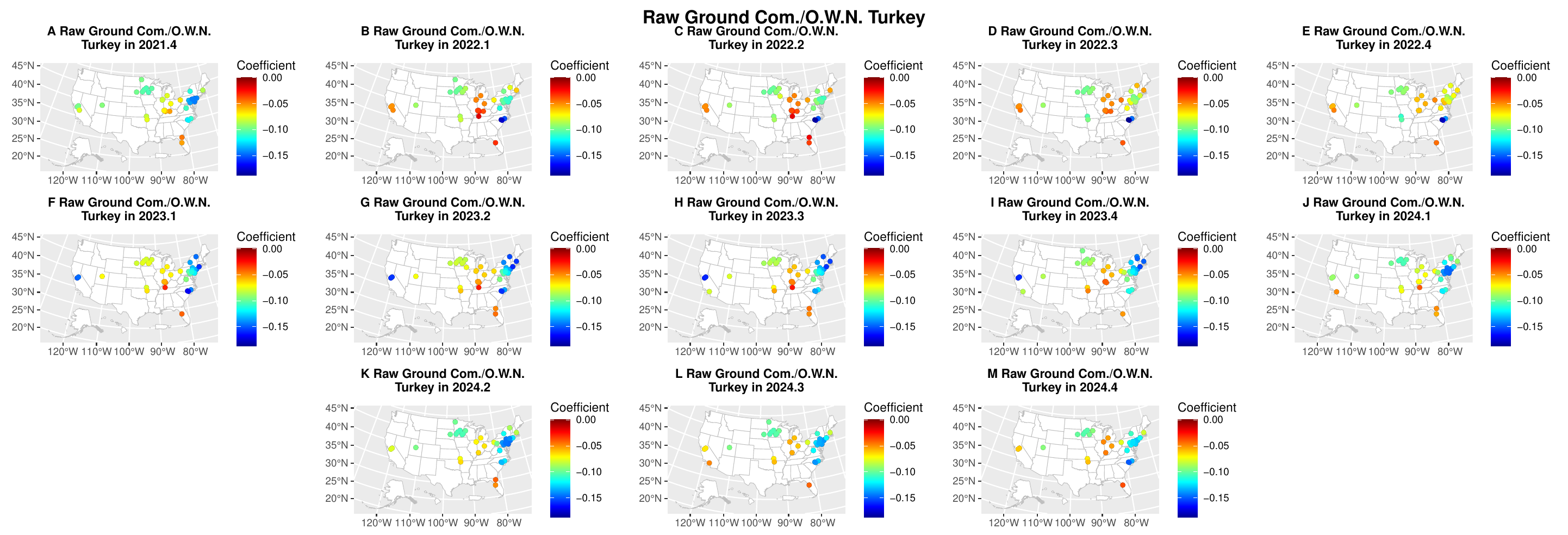

### S1n.pdf-1.png

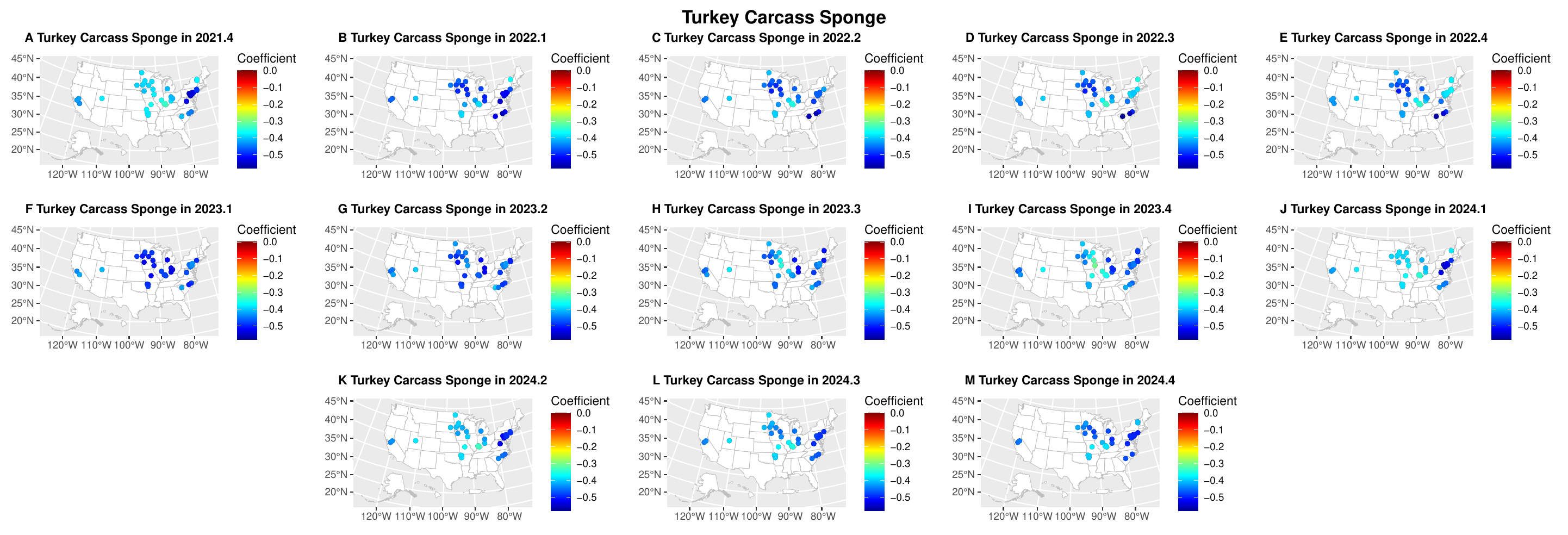

### Supplementary File 2

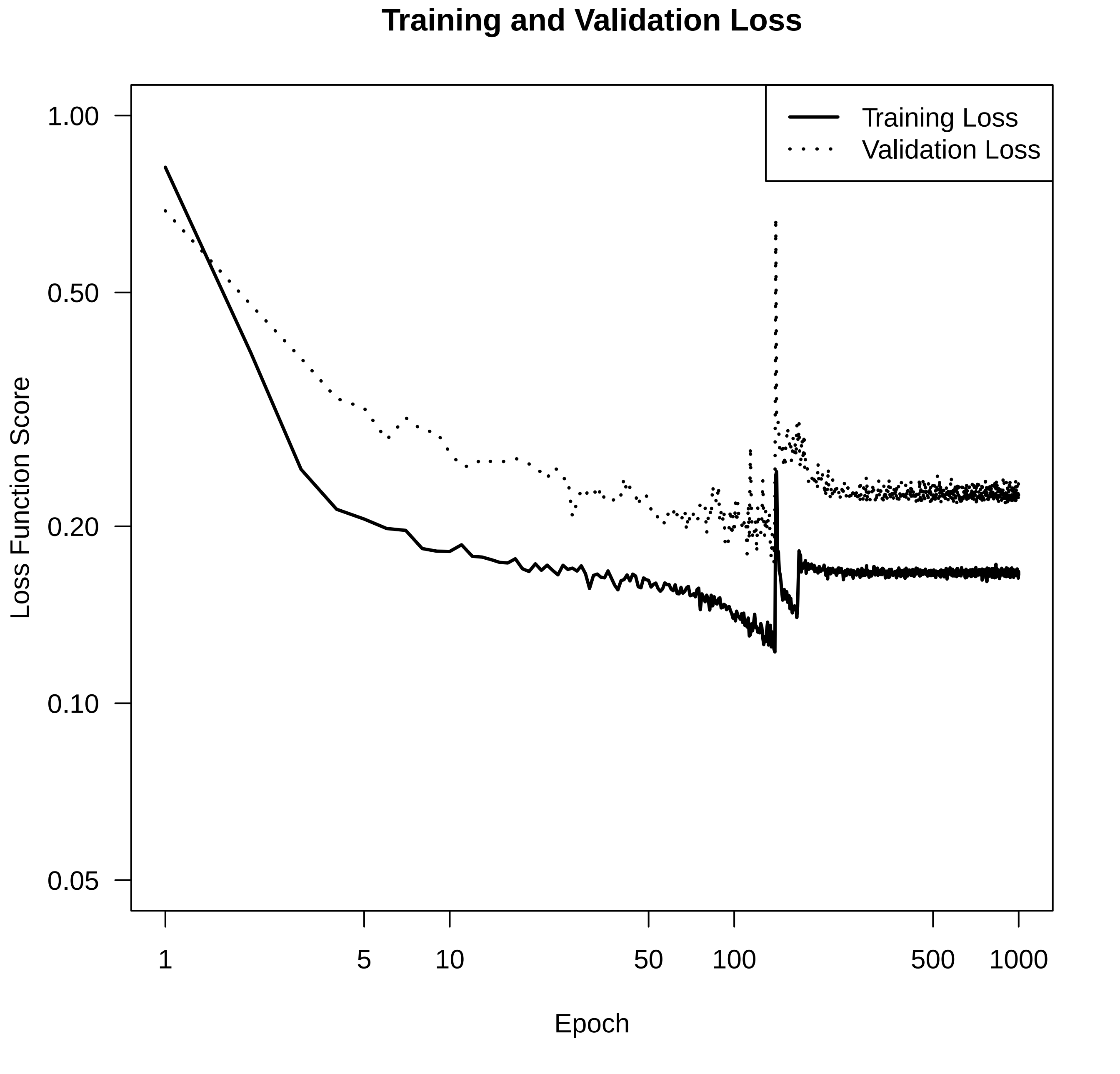
